# COVID-19; Systematic and literature review of transmission, case definitions, clinical management and clinical trials

**DOI:** 10.1101/2020.05.14.20102475

**Authors:** Laura McArthur, DhanaSekaran Sakthivel, Ricardo Ataide, Felicia Chan, Jack S. Richards, Charles A. Narh

## Abstract

**Background:** SARS-CoV-2, the viral agent responsible for coronavirus disease 2019 (COVID-19) was identified in Wuhan, China at the end of December 2019. It rapidly spread to the rest of the world, and was declared a Public Health Emergency of International Concern on the 30th of January 2020. Our understanding of the virus, it’s clinical manifestations and treatment options continues to evolve at an unparalleled pace.

**Objective:** This review sought to summarise the key literature regarding transmission, case definitions, clinical management and trials, and performed a systematic review of reported clinical data on COVID-19.

**Synthesis methods:** Two reviewers selected all the literature independently, and extracted information according to pre-defined topics.

**Results:** COVID-19 is pandemic with ∼4 million cases and 270,000 deaths in 210 countries as of 8 May 2020. Our review of reports showed that SARS-CoV-2 was mainly transmitted via inhalation of respiratory droplets containing the virus and had an incubation period of four to six days. The commonly reported symptoms were fever (80%) and cough (60%) across the spectrum of clinical disease – mild, moderate, severe and critical. Categorization of these cases for home care or hospital management need to be well defined considering the age of the patient and the presence of underlying co-morbidities. The case definitions we reviewed varied among affected countries, which could have contributed to the differences observed in the mean case fatality rates among continents – Oceania (1%), Asia (3%), Africa (4%), South America (5%), North America (6%) and Europe (10%). Asymptomatic cases, which constituted an estimated 80% of COVID-19 cases are a huge threat to control efforts.

**Conclusion:** The presence of fever and cough may be sufficient to warrant a COVID-19 testing but using these symptoms in isolation will miss a proportion of cases. A clear definition of a COVID-19 case is important for managing, treating and tracking clinical illness. While several treatments are in development or in clinical trials for COVID-19, home care of mild/moderate cases and hospital care for severe and critical cases remain the recommended management for the disease. Quarantine measures and social distancing can help control the spread of SARS-CoV-2.

## Introduction

In December 2019 a number of Chinese Health Centres reported a cluster of patients with pneumonia of unknown aetiology in Wuhan, Hubei Province (1, 2). Their clinical presentations were similar to the Severe Acute Respiratory Syndrome (SARS) outbreak that occurred in 2003 (3–5). COVID-19 is the third coronavirus disease to cause public health outbreaks and has spread more rapidly and widely than SARS and Middle East Respiratory Syndrome (MERS). As of 8^th^ May 2020, there were ∼4 million confirmed cases and ∼270,000 deaths in 210 countries and territories. This review provides a discussion of the disease transmission, clinical presentations, variability of case definitions and review of the clinical management and clinical trials of drugs for COVID-19.

## Methods

### Literature search

This study was conducted following the PRISMA recommended guidelines (see Supplementary data for PRISMA check list version 2009) where relevant. Literature search for articles relevant for the topics discussed in this review was performed using OVID Medline, PubMed and Google Scholar. The search terms included “SARS-CoV-2 and Wuhan”, “Transmission and reproductive number and SARS-CoV-2”, “SARS-CoV-2 and COVID-19”, “COVID-19 and case definitions”, “COVID-19 and symptoms and clinical”, “COVID-19 and asymptomatic”, “COVID-19 and risk factors”, “COVID-19 and treatments and hospital” and “COVID-19 and adults or children”. Furthermore, references within the articles that were returned by the search engines were also retrieved and reviewed if they contained relevant information for this current review. Medical review sites including BMJ Best Practice and UpToDate were reviewed periodically to provide background and to get updates on COVID-19. For case definitions, the WHO criteria and those published by individual countries in the six continents were obtained. For case management, the NIH Clinical Trials database (https://clinicaltrials.gov/) was reviewed. The list of drugs and treatments in development and clinical trials for COVID-19 were obtained through periodic searches of News articles, Press releases and reports in the medical journals. Relevant Preprints on biorxiv and Medrxiv were included only affect they were critically evaluated by two independent reviewers.

### Data collection and analysis

To determine the commonly reported symptoms for COVID-19, data was extracted from 15 studies that involved COVID-19 patients including children and adults (6–20) (Figure S1). For each study, the proportion of patients who reported or were observed to have a particular symptom was recorded in an excel table. For each symptom, the average proportion of the number of patients with that symptom was estimated and plotted with the upper standard error using Microsoft Excel. For each country, data on the total number of cases and deaths reported since the outbreak began till 8^th^ May 2020 were retrieved from Worldometer (21). The case fatality rate was estimated as a percentage of the quotient of the total number of deaths reported and total number of cases reported for each country. The total number of deaths and case fatality rates were plotted for countries with ≥ 15 deaths reported. All mapping analyses were performed using QGIS 3.12.1-București (http://qgis.osgeo.org). Shapefiles for country borders were downloaded from Natural Earth (Free vector and raster map data at naturalearthdata.com). The data extracted from Worldometer (21) was imported to Stata/SE 16.1 (StataCorp, TX, USA) and the country names were spelt to match those of the country shapefile. The resulting database was exported as a csv file and imported to QGIS where a join was performed using the MMQIS Plugin developed by Michael Minn v.2020.1.16. (http://michaelminn.com/linux/mmqgis/).

#### 1. Human-to-human transmission routes of SARS-CoV-2

SARS-CoV-2 is transmitted between humans via respiratory droplets which are produced when an infected individual talks, sneezes or coughs (Figure 1). Droplet transmission can occur within 1–4 meters (15, 22, 23). SARS-CoV-2 has been shown to survive in aerosolised form for more than 3 hours under experimental conditions, but this mechanical generation of aerosols is unlikely to mimic the true clinic scenario (24). Certain clinical procedures involving the upper airway, such as obtaining a nose or throat swab, endotracheal intubation, manual ventilation or nebulisation are capable of generating particles <5µm, allowing for airborne transmission in healthcare settings (23). In particular, intensive care units (ICU) have been associated with a higher risk of infection (22).

Fomite transmission, transmission from contact with contaminated surfaces, is possible with high rates of contamination of floors and the soles of healthcare staff as well as computer mice, doorknobs and trash cans (22). The virus is viable for up to 72 hours on plastic and stainless steel, 24 hours on cardboard and 4 hours on copper. These survival times appear to be longer than that of SARS-CoV under similar conditions and may contribute to the wider spread of SARS-CoV-2 than SARS-CoV (24).

Infection from direct contact with body fluids from infected individuals is likely to be another possible route of transmission. SARS-CoV-2 has been detected in saliva, blood, urine, tears, faeces and cerebrospinal fluid samples (25–29). While documented evidence of transmission through these alternate sources remains unsubstantiated, precautions when handling samples collected from suspected or confirmed cases is advisable.

#### 2. Rate of transmission of SARS-CoV-2

The basic reproductive number (R_0_) of SARS-CoV-2 varies between populations and depends on demographic and environmental factors as well as on the control interventions in place (30). At the onset of the outbreak in Wuhan, before travel restrictions were introduced, the estimated R_0_ was 2.35 (95% CI 1.15–4.77) but was reportedly reduced to 1.05 (95% CI 0.41–2.39) a week later (31). Similarly, aboard the Diamond Princess cruise ship – which carried ∼3600 people, had 712 cases of COVID-19 and 13 deaths reported – the R_0_ was reported as 14.8 before quarantine/isolation measures were introduced and was reduced to 1.78 after those measures were introduced (32). In closed settings, such as nursing facilities, SARS-CoV-2 may spread rapidly with one study finding a 64% positivity rate among residents 23 days after the first positive test (33). Of these patients, 56% tested positive while still asymptomatic, and it has been hypothesised that asymptomatic individuals contribute significantly to transmission (33).

#### 3. Clinical presentation of SARS-CoV-2 infections

The reported incubation period for SARS-CoV-2 has been variable between different studies, but has generally ranged between 2–11 days, with an average of 4–6 days (34). In one study, the incubation period was estimated at 4.9 days (95% CI 4.4–5.5) and was not significantly different from that of SARS-CoV (4.7, 95% CI 4.3–5.1) and MERS-CoV (5.8, 95% CI 5.0–6.5) (35).

Symptomatic patients with COVID-19 develop a clinical syndrome similar to the influenza (Figure 2). Our analysis of 15 studies involving COVID-19 patients showed that the majority of patients with clinical disease had fever (80%) and cough (60%) and myalgia/fatigue (40%; Figure 2). Other reported symptoms were less common and included dyspnoea, sore throat, diarrhoea, vomiting, pharyngeal congestion, headache, sputum production, anorexia and chest pain (1, 36–39). The majority of these symptoms appeared within 11 days post-infection but may vary depending on age and co-morbidities (6, 30, 40, 41). See section 6 for a review of factors associated with disease severity. See Figure S1 for a detailed analysis of the reported symptoms for each of the 15 studies were the clinical data in Figure 2 was obtained.

**Figure 1:**
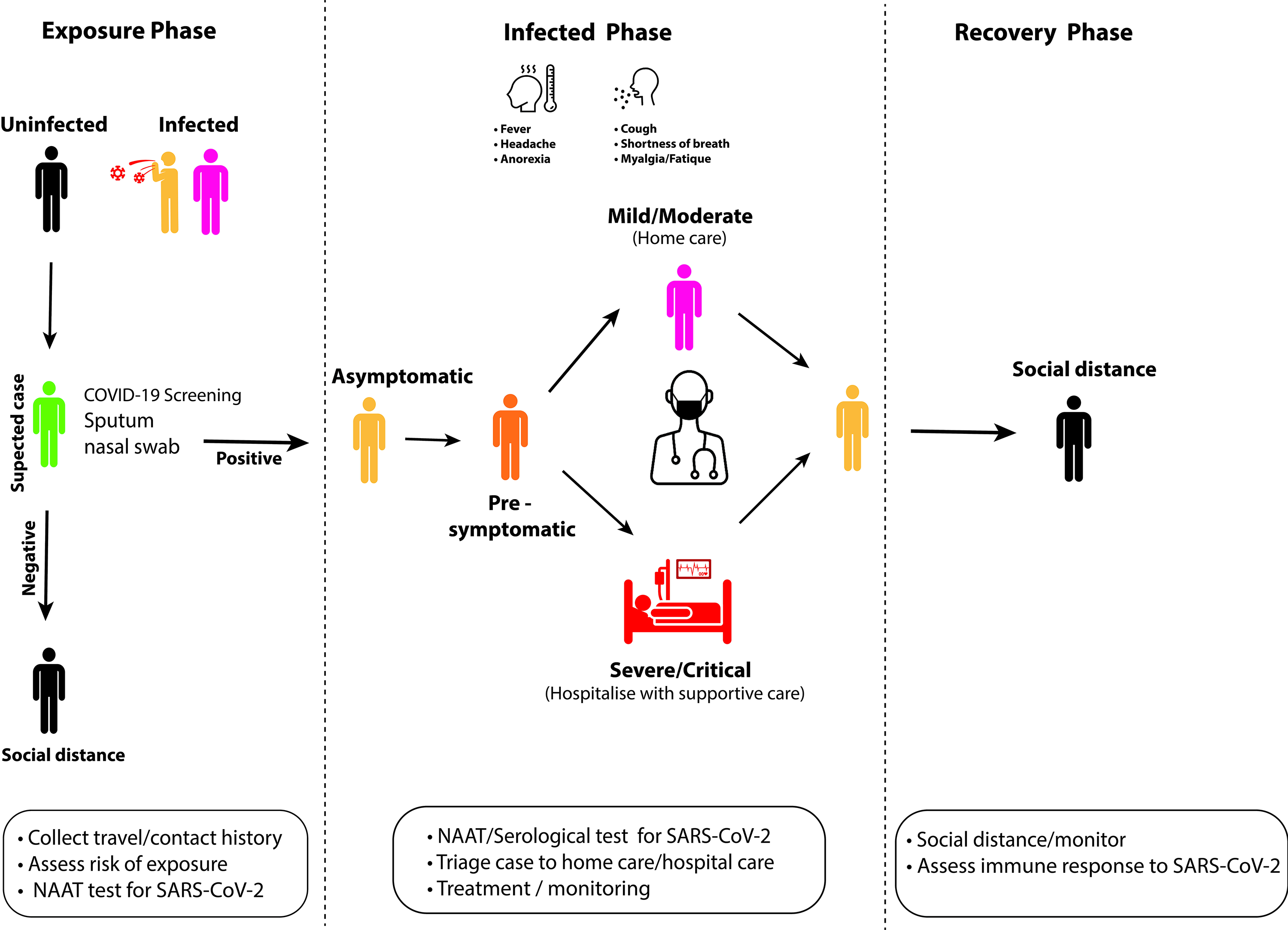
Identification and management of COVID-19 cases. Monitoring for suspected cases of COVID-19 is crucial to halt transmission of SARS-CoV-2. Suspected cases who have had contact with an infected person (asymptomatic/symptomatic) need to be isolated and screened for SARS-CoV-2 using sensitive nucleic acid amplification tests(NAATs). It is recommended that infected individuals who are asymptomatic self-isolate and be monitored at home. Individuals who progress to develop clinical disease can be triaged into mild/moderate and severe/critical case for clinical management/treatment. However, the presence of co-morbidities and the age of the patient can be used to triage patients for hospitalization or homecare. Once recovered, patients should be monitored since they could still be infectious.

**Figure 2:**
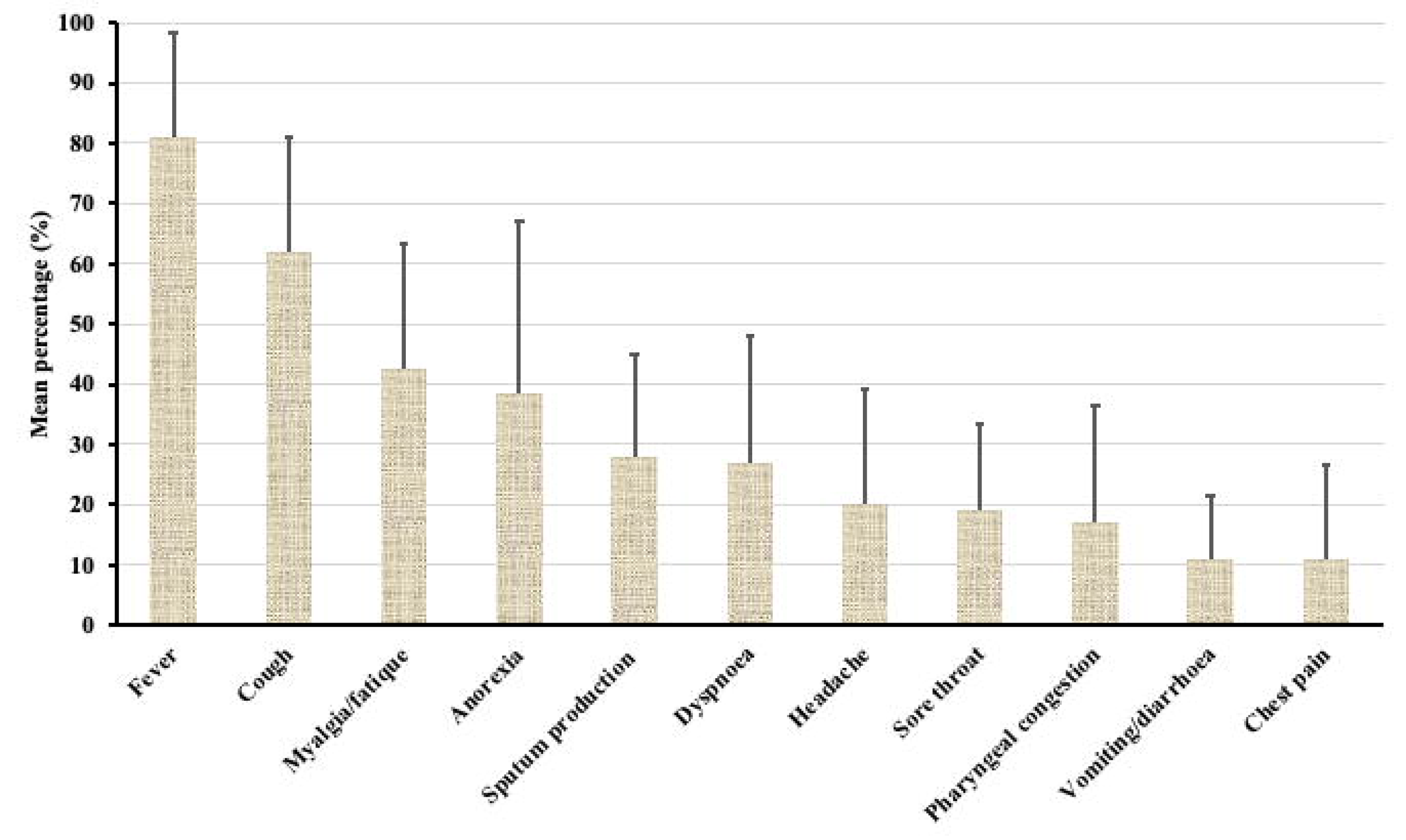
Commonly reported clinical symptoms of COVID-19. The data was obtained from 15 studies involving COVID-19 patients including children and adults. For the pooled analysis, the mean percentage of patients who developed a particular symptom was plotted with the upper standard error. Fever and cough were the commonly reported symptoms. Data was obtained from (6–20). A detailed analysis of reported clinical symptoms for each of the 15 studies is provided in Figure S1.

Where illness warrants hospitalisation, studies suggest that this usually occurs within five days of symptom onset (30), with a mean time to hospitalisation of seven days (7). The average time of hospital admission ranged from 12.8 – 17 days and was dependent on disease severity (6, 42). WHO recommends 2–6 weeks of hospitalisation to allow time for proper treatment and adequate recovery (43). The disease can progress from mild to critical with severe respiratory complications and multiple organ dysfunction, which can be fatal (1, 37, 44). In COVID-related deaths, the median time from symptom onset to death is about 13 days (45).

##### 3.1. Asymptomatic cases

Asymptomatic SARS-CoV-2 infections, where patients test positive for the virus yet remain clinically well, constitute a large proportion of COVID-19 cases. As such, a population screening strategy based on clinical symptoms alone will miss these cases. Estimates suggest that up to 80% of people who test positive for SARS-CoV-2 are asymptomatic (46). In a proportion of individuals who are initially asymptomatic, a clinical illness will subsequently develop, classifying this group as pre-symptomatic. A study of asymptomatic patients found that 67.3% had, as determined by computed tomography (CT), lung abnormalities at admission and 70.9% later developed a mild illness. In the 32.7% who did not have initial CT changes, only 11% later developed clinical illness (47). Pre-symptomatic cases have also comprised the majority of positive results when testing is performed in nursing facilities, with these cases likely contributing to transmission (33).

The distribution of asymptomatic cases varies with age, with the majority of infected children more likely to be asymptomatic compared to adults (48, 49). In an analysis of cruise ship infections and Japanese evacuees, mostly adults from Wuhan, asymptomatic cases were estimated to account for 17.9%-33.3% of the total SARS-CoV-2 infections (50). In recent reports from naval officials regarding an outbreak aboard the Theodore Roosevelt, approximately 60% of positive patients were asymptomatic at the time of testing (51). Given that the majority of these individuals were relatively fit, young and healthy individuals, it suggests that the disease may be more likely to be asymptomatic in this cohort, while still contributing to transmission.

While the degree of infectivity of these asymptomatic patients compared to those with clinical illness remains uncertain, there are documented cases of asymptomatic individuals transmitting the virus during the incubation period, and continuing to shed virus while convalescing (52). Numerous cases of pre-symptomatic transmission have been reported including data that suggested that 12.6% of cases in China were transmitted asymptomatically (53). These findings raised questions as to whether the majority of the patients who were classified as ‘pre-symptomatic’ may have had mild symptoms that went unreported (52, 54, 55). In Singapore, the CDC reports on seven clusters of patients where 6.4% of cases occurred through pre-symptomatic transmission, between 1–3 days.

##### 3.2. Mild illness

Mild COVID-19 illness is defined by uncomplicated symptoms which can be safely managed in the outpatient setting. While the World Health Organisation separates this category into those with and without a mild pneumonia, this distinction does not influence management and is difficult to make in patients managed outside of hospital. For the purposes of this review, all cases suitable for management in an outpatient setting will be considered together.

Mild COVID-19 is perhaps less well understood than more severe disease phenotypes as it is believed that the majority of such cases do not present for testing. Based on current data, approximately 81% of confirmed SARS-CoV-2 infections are regarded as mild (26); however, it is likely that the number of mild cases has been significantly underestimated. During the early period of the disease outbreak in China, it was estimated that 86% of infections went undocumented due to the fact that those infected developed non-severe symptoms and therefore did not present for testing (56). The study further estimated that undocumented infections were the sources of infection of 79% of all documented infections (57).

Symptoms of mild COVID-19 are classically of an upper respiratory tract infection, with atypical presentations being more common in elderly and immunosuppressed individuals (58). It appears that fever is less characteristic of mild cases. In a Dutch study of patients identified through screening, 53.5% had fever compared to 77–98.6% of hospitalised patients. Headache and pharyngeal congestion were comparatively prominent with malaise, cough and myalgia being less typical than in severe cases (7, 11, 12). Although fatalities have been reported among children, they seem more likely to experience a mild illness and the case fatality rate appears significantly lower compared to adults (59). As children can still contribute to transmission, considerations for social distancing remain relevant despite the generally comparative mild illness phenotype in this cohort (41).

##### 3.3. Severe and critical illness

Severe COVID-19 is defined by symptoms of significant respiratory distress which in adults are tachypnoea ≥ 30 breaths per minute; oxygen saturation ≤ 93%; PaO_2_/FiO_2_ ratio < 300mmHg; lung infiltrates > 50% within 24–48 hours; or clinical assessment of severe distress (58, 60). This definition encompasses Acute Respiratory Distress Syndrome (ARDS), defined by acute onset, PaO_2_/FiO_2_ ratio and bilateral infiltrates on CXR (61). Through the course of the illness, most patients will develop a fever, 77–98.6%, and cough, 48.2–76% (6, 8, 26, 47).

Other common symptoms include myalgia or fatigue, which appear early in the illness and seen in around 18–32.1% of cases (8), sore throat (19) and dyspnoea (26). Fatigue may ultimately occur in up to 69.6% of patients through the course of the illness but it is non-specific (47). Anosmia has been reported as a commonly observed symptom and in a recent Italian study 33.9% of patients reported a disturbance of smell or taste, with most experiencing this prior to their admission to hospital. Less common are gastrointestinal symptoms such as diarrhoea and vomiting, as well as chest pain and pharyngeal congestion. However, it is worth noting that while some symptoms are common – with fever in particular being regarded as a defining symptom of the illness – they are by no means necessary and will not always be present throughout the course of the disease.

Severe COVID-19 occurred in 14% of documented SARS-CoV-2 infections in China during the initial epidemic period (62). Further studies have determined a need for non-invasive ventilation consistent with severe COVID-19 in 10.8% (7) to 13.1% (63) of admitted patients. In addition, current literature records Acute Respiratory Distress Syndrome (ARDS) in 3.4% (6), 17% (56), 29% (26) and 41.8% (64) of hospitalised COVID-19 patients. Where ARDS develops, Wang et al. observed a mean time from first symptoms to ARDS to be 8 days (7).

The majority of patients, 70.3–83.2%, with severe cases develop lymphocytopenia (6–8) and CT changes have been identified in 56% of patients on admission to hospital (6, 65). CT changes are of particular note as these may be identified even in asymptomatic individuals, and if present increase the likelihood of a clinical illness developing (47). A moderate disease phenotype may exist in which patients do not meet criteria for severe or critical disease but nevertheless require in-hospital management. This is commonly due to comorbidities, inability to tolerate oral intake, or respiratory distress not yet meeting thresholds for severe disease.

Severe COVID-19 appears to be more common in men, with 54.3 – 68% of hospitalised patients in studies being male (47, 56) and asymptomatic or mild illnesses having an apparent female preponderance (47). Severe disease has likewise been associated with comorbidities albeit severe illness and death can occur in previously young, healthy individuals including infants (63, 66).

Critical cases of COVID-19 are defined by respiratory failure requiring mechanical ventilation; septic shock; or organ dysfunction necessitating intensive care (60). Critical COVID-19 cases were recorded in 5–6% of SARS-CoV-2 infections during the epidemic phase in China (60, 63). In other studies of hospitalised patients, the proportion of COVID-19 cases requiring intensive care unit (ICU) admission ranged from 5% to 32% (6, 7, 26, 67). A need for invasive mechanical ventilation has been reported in 2.3–12.3% of hospitalised patients (6, 7) and ultimate requirement for extracorporeal membranous oxygenation (ECMO) in 0.5–3% of patients (6, 56). Critical COVID-19 has a case fatality rate of 49.0% recorded among those with critical disease in China (63).

##### 3.4 Recovery from COVID-19

Once patients have recovered, there is some suggestion they may still be able to transmit the virus, with case reports of positive RT-PCR throat swabs 5–13 days after symptom resolution, and potentially present in patients who have previously been swab-negative for SARS-CoV-2 (68).

Viral shedding has been observed for up to 37 days in survivors with a median duration of 20 days (69). SARS-CoV-2 has also been detected in faecal specimens 18–30 days after illness onset in children (19), suggesting that prolonged shedding may still be possible during and after recovery.

To date, studies of vertical transmission of SARS-CoV-2 remain limited but as of yet there are no definitive documented cases (70–72). While SARS-CoV-2 specific IgM has been detected in infant sera of COVID-19 positive mothers, it is as yet uncertain whether this represents passive immunization, such as through an altered placenta or disease transmission with infant immune response (73).

#### 4. COVID-19 case definitions

Defining the scope of the COVID-19 pandemic has been affected by the need to refine case definitions as the pandemic progressed and as clinical presentations become more clearly defined. These case definitions have been used to determine whom to test and to guide case investigations of possible contacts. By influencing testing algorithms, they have greatly impacted the confirmed test outcomes. As an example, case definitions in China were changed on the 12^th^ February 2020, to include clinically-diagnosed mild cases, resulting in an increase of > 15,000 in a single day (74). Thus, developing consistent case definitions, whenever possible, is necessary to track metrics of the disease and its spread.

COVID-19 case definitions have been developed and modified in different jurisdictions according to local circumstances and authorities (Table 1). In Canada, the definition of a probable case has been widened to require only one symptom of illness in addition to an epidemiological risk factor; and the definition of such a risk factor has also been expanded to encompass other sources of exposure, such as laboratory materials. In Australia, where case definitions are used to determine testing priorities, additional efforts have been devoted to defining high-risk settings such as residential facilities and the nature of a close contact as it relates to in-person interactions and proximity (Table 1). These definitions are highly significant as they determine who receives testing and which patients need to be regarded as at-risk for transmitting the virus. These precautions require resources – including equipment and personnel – such that case definitions must balance capturing possible COVID-19 infections against burdening the healthcare system with individuals with a low probability of infection.

**Table 1:**
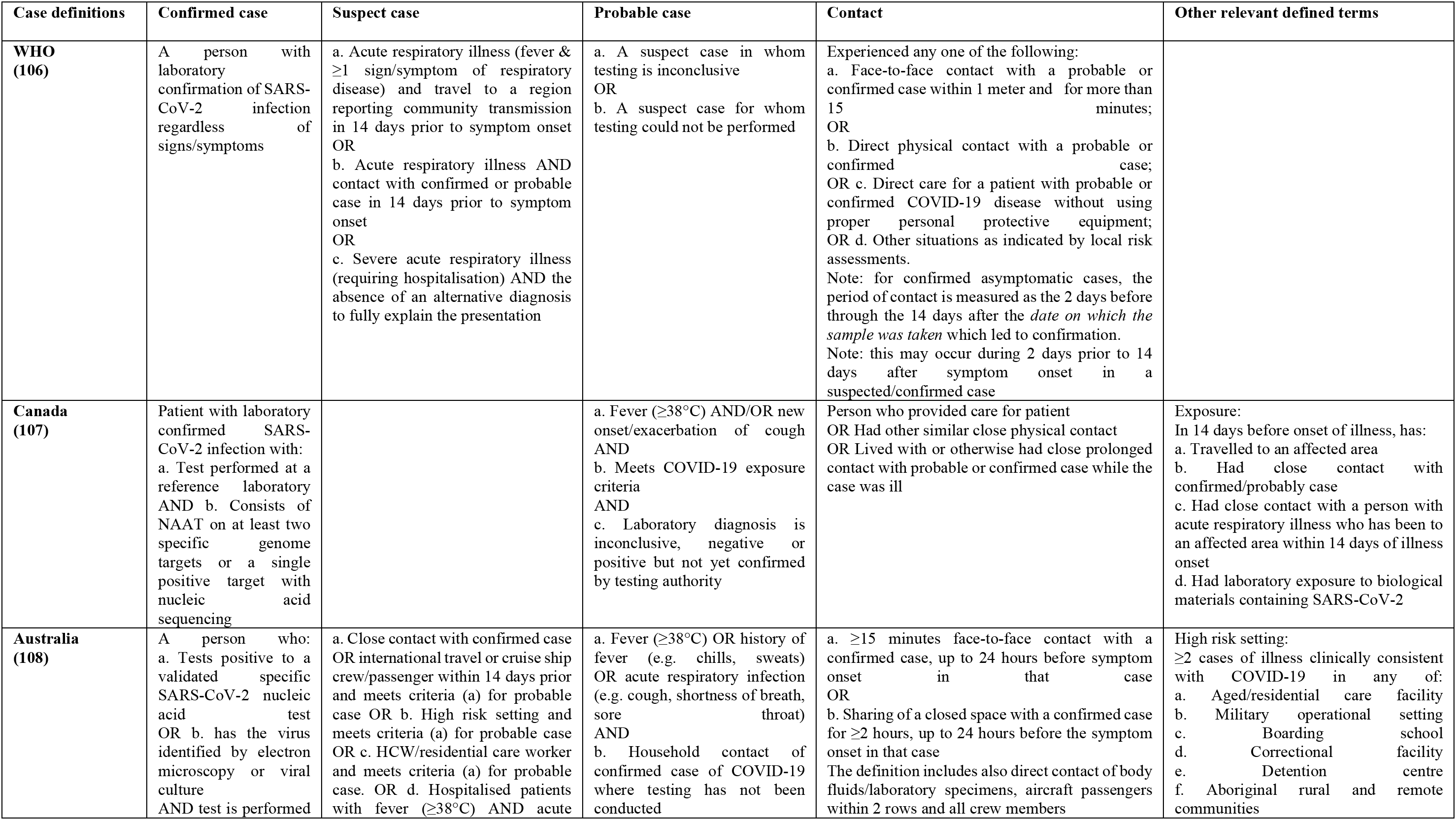
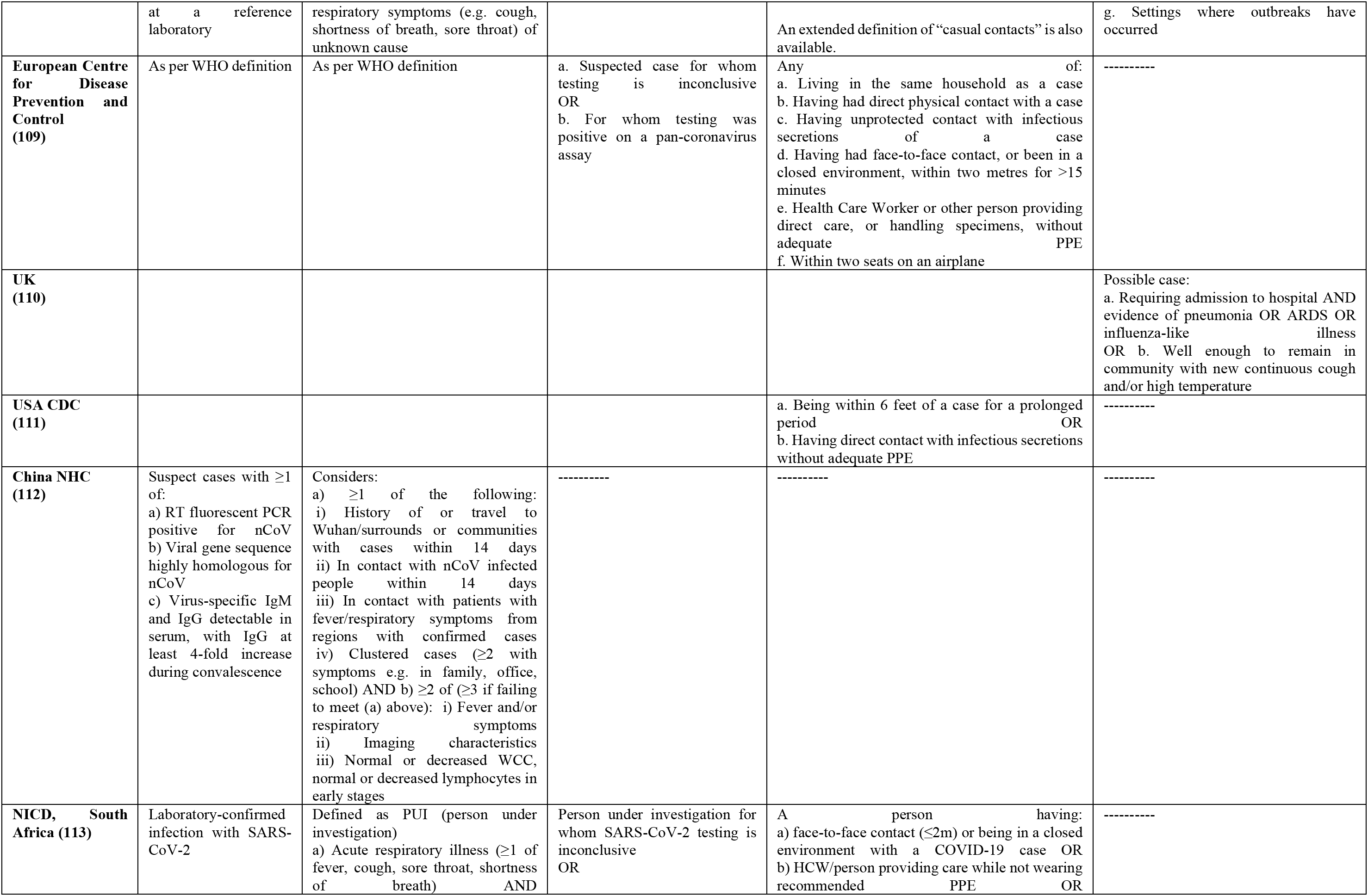
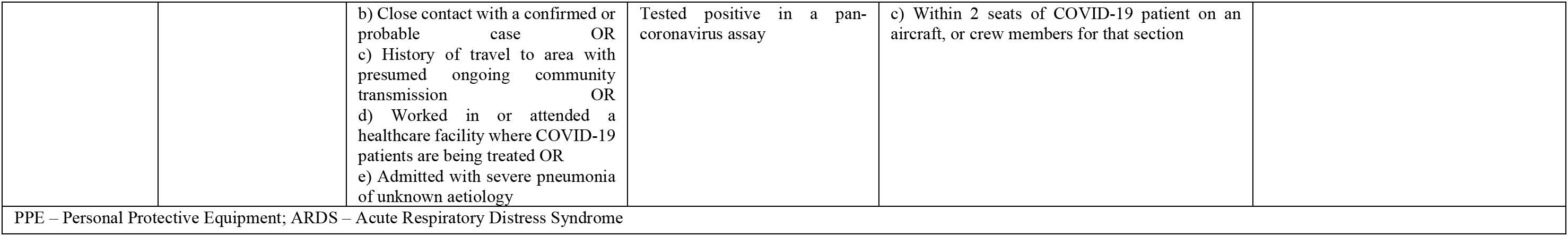
Case definitions of COVID-19

**Table 2:**
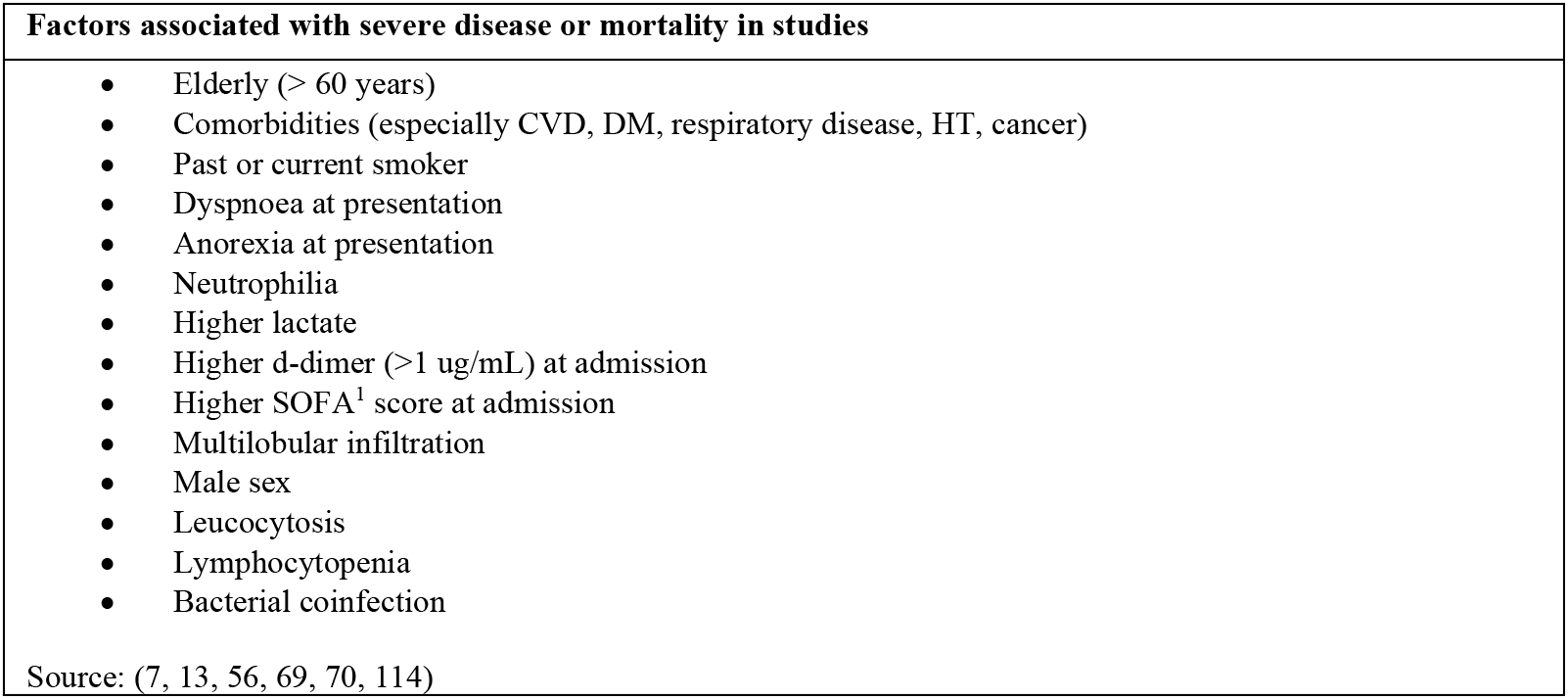
Factors associated with severe COVID-19 and mortality

#### 5. Burden and case fatality of COVID-19

As of 8^th^ May 2020, ∼4 million confirmed cases and 270,000 deaths due to COVID-19 were reported from 210 countries and territories (Figure 3A) (21). In Asia, where the epidemic started, ∼620,000 cases and ∼21,200 deaths were reported with estimated case mean case fatality of 3%. Although the hardest hit countries in this region were Turkey (∼134,000 cases: ∼3,600 deaths; 3% fatality rate), Iran (∼103,000; ∼6,500; 6%) and China (∼82, 900; ∼4,600; 6%), Indonesia and the Philippines had the highest case fatality, ∼7% (Figure 3B). It is worth noting that the case definitions were not the same which may affect testing and management of cases. As the epidemic spread farther outside Asia, Europe became the hotspot with ∼1.6 million cases and ∼150, 000 deaths with a mean case fatality of 10% (Figure 3B). The five hardest hit countries include Spain (∼256,900; ∼26,100; 10%), Italy (∼215,900; ∼30,000; 14%), UK (∼206,700; ∼30,600; 15%), Russia (∼177,200; 1,600; 1%) and France (∼174,800; 26,000; 15%). In North America (∼1,412,000; ∼85,200; 6%), the United States (∼1,300,000; ∼76,900; 6%) was the hotspot, contributing ∼92% of the total cases from the region (Figure 3A). In South America (∼267,900; 13,600; 5%), Brazil (∼135,800; ∼9,200; 7%), Peru (∼58,500; ∼1,600; 3%) and Ecuador (∼30,300; ∼1,600; 5%) were the hardest hit (Figure 3A). In Africa, where ∼55,400 cases were confirmed and ∼2,100 deaths occurred with a mean case fatality of 4%, South Africa (∼8,200; 160; 2%), Egypt(∼8,000; 500; 6%), Morocco (5,500; 180; 3%) and Algeria (5,200; 500; 9%) were the hardest hit(Figure 3B). Oceania had the lowest number of cases reported so far with ∼8,500 cases and 120 deaths with a mean case fatality of 1%, with the hardest hit country being Australia (7,000; 100; 1%, Figure 3B). As the pandemic continues, these figures and trends in cases and deaths may change, and thus, it is important for countries to keep track to inform public health control efforts.

**Figure 3:**
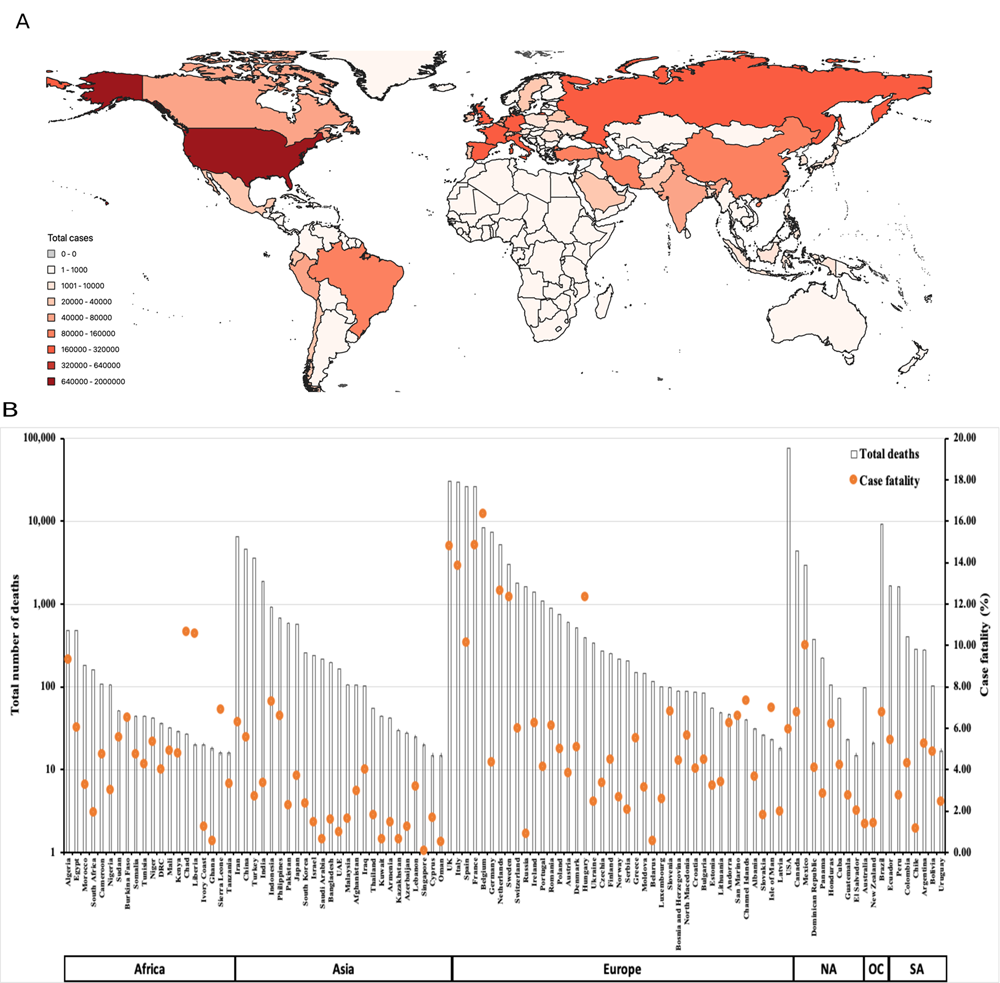
Global burden and case fatality rates of COVID-19. A: Total number of cases reported as of 8^th^ May 2020. B: Total number of deaths (primary axis) and estimated case fatality rates (secondary axis) for countries with ≥ 15 deaths reported as of 8^th^May 2020. Total number cases and deaths data were obtained from Worldometer(https://www.worldometers.info/coronavirus/) on 8^th^ May 2020. OC, Oceania, SA, South America, NA, North America.

In different healthcare settings, the case fatality may vary. Studies of hospitalised patients have reported varying fatality rates ranging from 1.4 to 14% (6, 41, 74). However, several of such studies are likely underestimating true fatality rates in these patient groups given that the percentage of patients recovered to discharge by the conclusion of the study often remains low – 31% in the analysis by Chen et al, where case fatality rate was found to be 11% (10) and 34.1% in the study by Wang et al, where case fatality rate was 4.3% (7). In studies that have used the formula of fatal cases/(fatal + recovered) to calculate fatality rates, these have been found to be higher, estimated at 14% among hospitalised patients (74).

However, when considering all cases of SARS-CoV infection, including those that are not hospitalised, it is likely that the vast majority of mild cases are undocumented and therefore not included in calculations of case fatality rate – which would render it significantly lower than previously believed (57). With these factors complicating estimations of case fatality rate, it is difficult to determine a precise risk, but recent estimates taking these factors into account put the case fatality rate at 0.51% (75). This is supported by data from cruise ship cohorts. Of cases from the Diamond Princess, a population in which many of the case-finding difficulties were eliminated, studies recorded the case fatality rate at 0.99% (76).

Despite this uncertainty regarding the precise case fatality rates, it does appear clear that there are several significant risk factors for fatal illness. In early analysis of Chinese data, the risk of death was higher with increasing age, climbing to 8% among 70–79 year-olds and 14.8% among those ≥80 years (62). This was supported by data from Italy (77) where the higher fatality rate was associated with an older population (with 23% of Italy being ≥65 years) and age-specific death rates rose to 20.2% among those ≥80 years and 22.7% among ≥90 years. Likewise, in some studies of hospitalised patients age was associated with disease severity where the risk of severe disease increased with age (6, 47, 60, 78).

#### 6. Factors associated with infection and morbidity

COVID-19 mortality risk has also been consistently associated with comorbidities (6, 8). In Wu’s analysis, it rose from 2.3% in the general population to 10.5% among those with cardiovascular diseases, 7.3% among patients with diabetes mellitus, 6.3% among those with chronic respiratory diseases, 6% among patients with hypertension and 5.6% in cancer patients (63). A subset of fatal cases from Italy have been reported in which 99.2% had comorbidities of some description, with 48.5% having at least three underlying diseases (77). In addition, Yang et al. identified chronic illness in 40% of patients who fell critically ill (13). Other predictors of severe illness and death may include presentation with dyspnoea (present in 59.5–63.9% of ARDS cases, versus 19.6-25.6% of less severe illness) and anorexia (66.7% vs. 30.4%) (7, 64). Severe illness has also been associated with smoking (6) and the presence of neutrophilia, higher lactate and d-dimer (64).

Interestingly, host genetic factors including polymorphisms in the human receptor for SARS-CoV-2, the angiotensin-converting enzyme 2 (ACE2), may play a role in infection and severity of COVID-19 (79–81). Compared to European populations, East Asian populations were found to have a higher allele frequency of the ACE2 variant, associated with higher expression of ACE2 receptors (82). This suggests that East Asian and China population might have an increased susceptibility to SARS-CoV-2 infection as compared to European individuals. In contrast, Cai (83) found no difference in the expression level of ACE2 between Asians and Caucasians. Among genders, a study by Zhou *et al*. (84) found that Chinese males expressed higher levels of ACE2 receptors. These findings are preliminary and inconclusive but they do warrant further investigations.

#### 7. Management of COVID-19 cases

For the majority of patients, COVID-19 presents as a mild illness that can be managed at home with rest and simple analgesics/antipyretics for symptom relief (Figure 1). Paracetamol has been suggested as the drug of choice for symptom relief while anecdotal evidence of NSAID-associated harm in COVID-19 infected patients is being investigated (85). Glucocorticoids, previously used in SARS, have been found to be harmful in influenza and MERS-CoV; the CDC and WHO currently recommend against their use for COVID-19 (58).

Currently, most healthcare facilities are attempting to manage milder cases of COVID-19 on an outpatient basis, with patients self-isolating in their own homes. However, it has been questioned whether early intervention with oxygen therapy may be beneficial, with doctors reporting cases of COVID-19 patients with extremely low oxygen saturations, despite relatively mild symptoms (86). These patients may be at increased risk of rapid deterioration and may benefit from early oxygen therapy (86).

Once patients are admitted to hospital, treatments fall into three key categories: supportive care; treatment of coinfection and comorbidity; and disease-modifying treatments, which currently remain experimental.

For patients with severe or critical SARS-CoV-2, supportive care is the current mainstay of treatment (Figure 1). This includes attention to fluids and electrolytes, monitoring for complications, facilitating symptomatic management and providing respiratory support. Respiratory support can be provided in a stepwise fashion as required, moving from oxygen therapy through to non-invasive ventilation then intubation and mechanical ventilation. In ARDS, there is some evidence that prone positioning of patients may improve oxygenation and it is currently being recommended for critical care of COVID-19 patients (59). Extracorporeal membranous oxygenation may be used if available for refractory hypoxia. It is important to note that while respiratory support measures are integral to COVID-19 management, they also create high-risk environments in which airborne transmission of the virus may be possible (23). These measures include intubation, nebulised treatments, moving patients to prone position and positive-pressure NIV. During these activities, health care personnel require personal protective equipment including N95 mask and eye protection.

Where COVID-19 is causing severe illness or sepsis, the World Health Organisation recommends empirical antimicrobial treatment, with other sources suggesting this be considered in any severe infection (59). In addition, consideration may be given to a neuraminidase inhibitor in the event of co-infection with influenza. Coinfection of COVID-19 patients with other pathogens may be common, occurring in 22% of cases in some reports (87). However, this depends on region and season. Coinfection with other respiratory pathogens may increase COVID-19 severity. As such, severe disease warrants testing and treatment.

#### 8. Drugs in development and in clinical trials for COVID-19

There are a range of drugs and treatments currently under investigation due to their potential to modify some aspect of the COVID-19 disease course (Table 3).

**Table 3:**
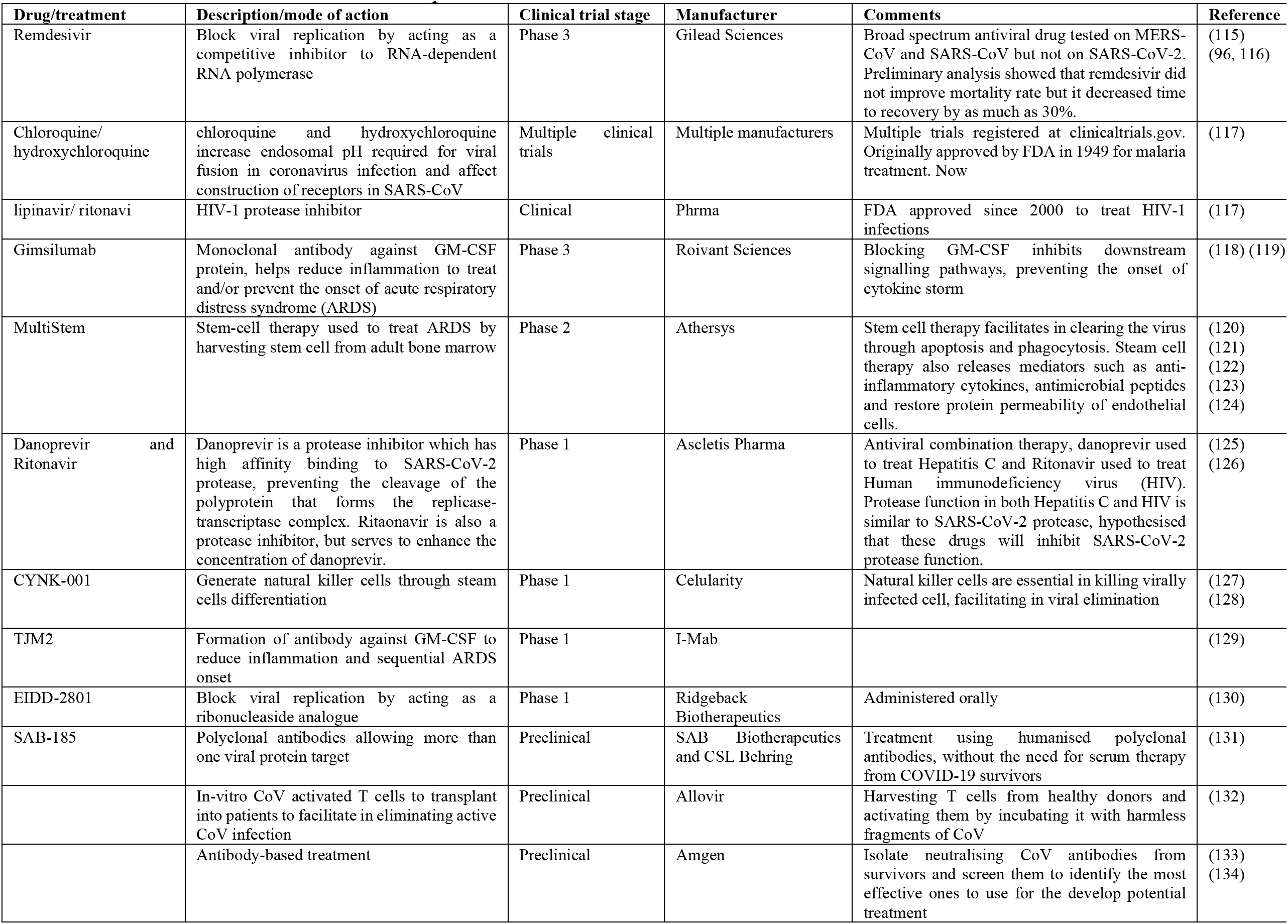
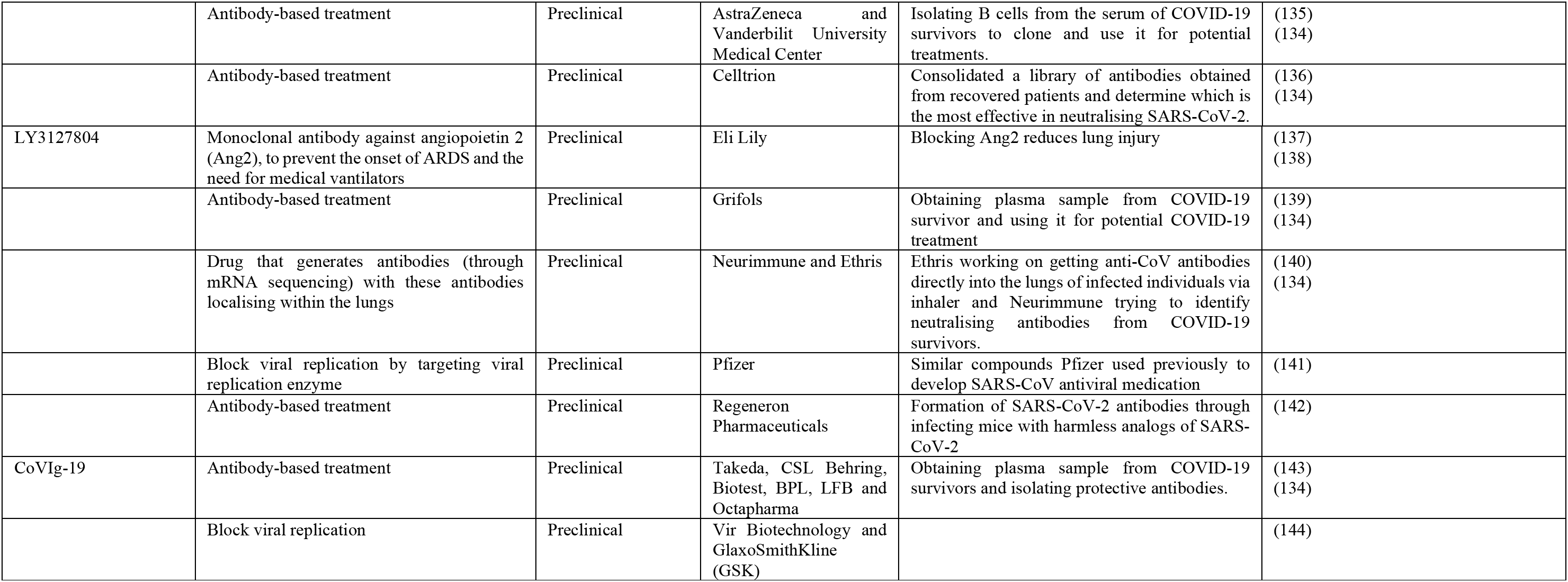
Current treatments in development and in clinical trials for COVID-19

##### Remdesivir (GS-5734)

An adenosine nucleoside analogue produced by Gilead Sciences, remdesivir has previously been under investigation for its use against RNA viruses including Ebola virus (88). Due to pre-existing evidence of efficacy against SARS-CoV and MERS-CoV in vitro and in animal models (89–92), it has been provided on compassionate grounds for some patients with COVID-19 (88) and has recently being approved by the US FDA for emergency use for the treatment of severe cases (93). Multiple trials have been launched into its effectiveness. There is currently limited evidence of its efficacy, with *in vitro* studies demonstrating inhibition of COVID-19 (94) and a US case report of a patient who improved following IV remedesivir administration (95). A non-randomised study of patients with severe disease who were treated with the drug, based on compassionate grounds, suggested that 68% experienced improvement in requirements for respiratory support (88). A recent report by US officials claimed that preliminary analysis showed that remdesivir did not improve mortality rate but it decreased time to recovery by as much as 30% (96). Multiple clinical trials into its efficacy are still in progress, including DisCoVeRy by INSERM (NCT04315948), ACTT through NIAID (NCT04280705) and studies through Capital Medical University (NCT04252664; NCT04257656) and Gilead (NCT04292730; NCT04292899).

##### Chloroquine and hydroxychloroquine

Traditionally used as anti-malarials, both chloroquine and hydroxychloroquine increase endosomal pH required for viral fusion in coronavirus infection and affect construction of receptors in SARS-CoV (94). There is some evidence that both drugs had efficacy against SARS-CoV and SARS-CoV-2 (94, 97, 98). Both drugs have a long history of use and a well-established adverse effect profile; they have been suggested for use in treating COVID-19. A State Council of China news briefing in February 2020 reported that chloroquine phosphate was superior in a treatment arm as compared to controls in improving clinical outcomes (99). An expert consensus supporting its use has been released (100) but its efficacy has yet to be conclusively demonstrated in clinical trials. A non-randomised control trial in France found that hydroxychloroquine reduced nasal viral load six days post-treatment and had high efficacy when combined with azithromycin. But there was no correlation with reported clinical outcomes (101). In a randomised trial of hydroxychloroquine among hospitalised patients, the drug improved clinical symptoms when compared to standard of care but did not improve negative conversion rate of SARS-CoV-2 (102). In another trial involving hospitalised patients in China (currently awaiting peer-review), the drug reduced the time to recovery from fever, cough and pneumonia (18). More recently, observational studies involving 1,376 patients with COVID-19 in the United States showed no significant association between hydroxychloroquine use for severe illness and intubation or death (103).

Multiple trials are currently underway to assess the efficacy of these drugs for COVID-19 treatment. This include THDMS-COVID19 through Rajavithi hospital (NCT04303299), Alliance Covid-19 Brasil II through the Hospital Israelita Alberta Einstein (NCT04321278), NO COVID-19 through University hospital, Akershus (NCT04316377) and HCQ4COV19 through Dundacio Lluita Contra la DIDA (NCT04304053).

##### Ritonavir/lopinavir +/− interferon beta

Ritonavir and lopinavir are protease inhibitors used in combination treatment for HIV. A study suggested that a combination therapy of the two drugs is effective against MERS in animal models (90). This combination is being trialled in COVID-19 patients (104). While an early case report showed that treatment with this therapy lead to a decrease in the viral load (105), a randomised trial of hospitalised patients with severe disease observed no benefit of the antiviral combination compared to standard of care (27).

On an ongoing basis, further studies are being conducted, including THDMS-COVID19 through Rajavithi hospital (NCT04303299), CORIPREV-LR (NCT04321174) and studies being conducted through the Asan Medical Centre (NCT04307693), first Affiliated Hospital of Zhejiang University (NCT04261907) and Tongji Hospital (NCT04255017). In addition, this combination is being trialled in patients outside of study settings (104) and is also being used as the standard of care to which new treatments are being compared.

These four medications/combinations – remdesivir, lopinavir/ritonavir, lopinavir/ritonavir + interferon beta and chloroquine – have been incorporated into the recently announced WHO megatrial (the “Solidarity Trial”) – an international collaboration investigating COVID-19 treatments.

#### 9. Conclusion

SARS-CoV-2, the causative agent of the novel coronavirus disease, COVID-19, has infected ∼4 million people and caused ∼140,000 deaths globally. A clear description of symptoms associated with the disease is crucial to define the case definition for clinical management and for epidemiological purposes. The majority of patients are likely to remain asymptomatic or mildly symptomatic. Those that do develop clinical disease will usually have fever and cough. The presence of either of these symptoms should be a basis for suspecting a SARS-CoV-2 infection. However, since other respiratory viruses present with similar symptoms, a laboratory test specific to SARS-CoV-2 should be performed.

Nearly, every country in the world has been hit by this pandemic. It is important to note that the definitions for COVID-19 cases vary slightly between countries and territories affected by the disease and this in turn may have affected the public health response. When defining cases, it is important that national guidelines factor in the presence of co-morbidities including cardiovascular diseases, diabetes and cancer, which increase the risk of developing severe and/or critical disease and increase the risk of fatality. Where transmission is still in the exponential phase of SARS-CoV-2 infections, it is critical that clinical cases are triaged to prioritize management and treatment without overwhelming the healthcare system.

Several drugs are in clinical trials but it may take months before any become available for clinical use. Hence, it is important that quarantine and isolation measures are strictly enforced to control the disease outbreak. A major challenge though to controlling SARS-CoV-2 transmission is how to identify the “silent spreaders” who are asymptomatic carriers of the infection.

### Limitations of the study

During the review period, the data on COVID-19 constantly changed with increasing amounts of literature both peer-reviewed and non-peer-reviewed. COVID-19 data was dependent on country level definitions and testing rates.

## Data Availability

All data have been included in the manuscript.

## Author Contributions

CAN conceived and designed the study. CAN and LM performed the majority of the writing. DS, RA and JSR contributed to the writing. CAN, LM, DS and FC performed the literature review and data collection. CAN and RA performed the data analysis. RA and JSR critically revised the manuscript. All authors read and approved the manuscript for publication.

## Funding

This work was supported by the National Health and Medical Research Council (NHMRC) of Australia [APP1161076 to J.S.R.]. Burnet Institute received funding from the NHMRC Independent Research Institutes Infrastructure Support Scheme, and the Victorian State Government Operational Infrastructure Support Scheme. The funders had no role in study design, data collection and analysis, decision to publish, or preparation of the manuscript.

**Figure S1:**
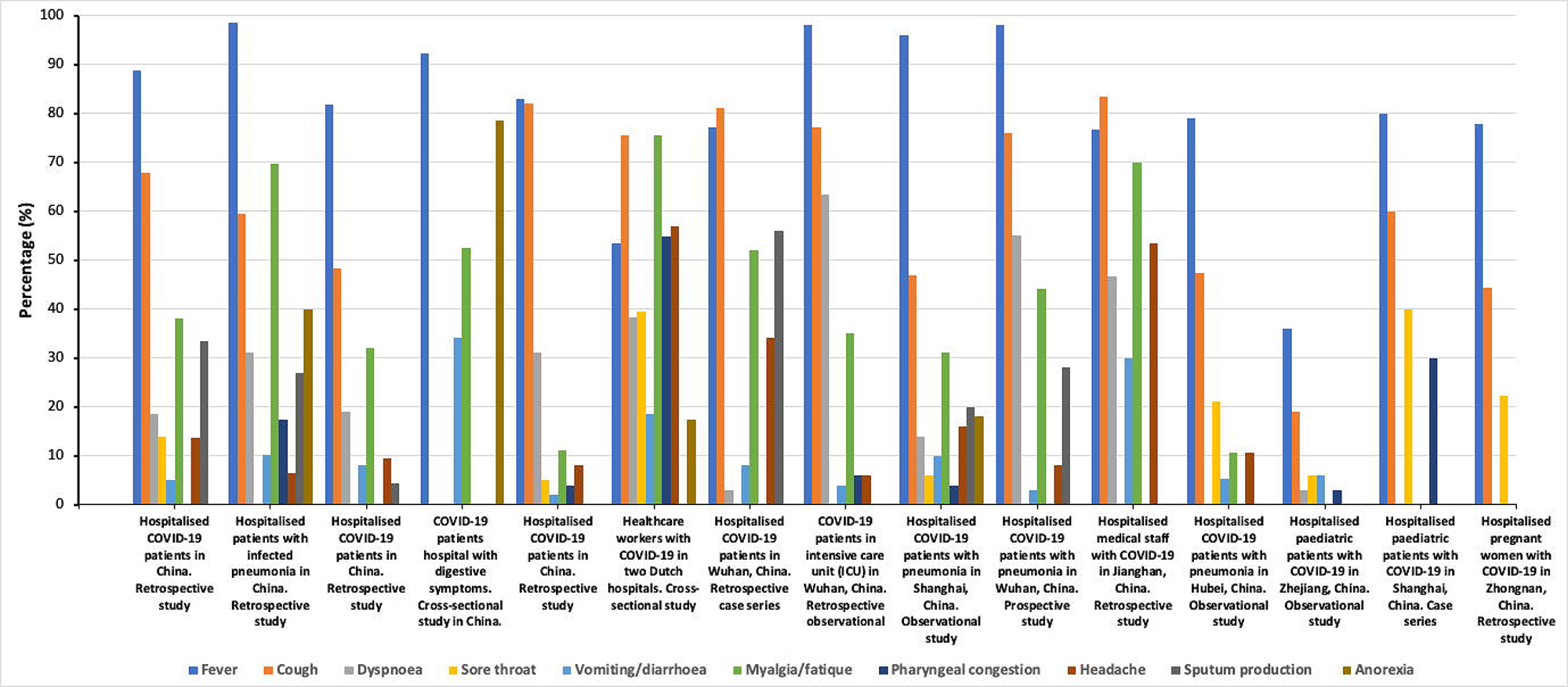
Commonly reported COVID-19 clinical symptoms in 15 studies. For each study, the percentage of patients who developed a particular symptom was plotted. Fever and cough were the commonly reported symptoms. Data was obtained from (6–20).

